## Supplement for "Completeness and consistency of ethnicity categorisations in administrative data: A comparison between linked health and education datasets"

**Supplement 1: The RECORD statement – checklist of items, extended from the STROBE statement, that should be reported in observational studies using routinely collected health data. Items specific to cross-sectional/case-control studies have been removed.**

|  | **Item No.** | **STROBE items** | **Page** | **RECORD items** | **Page** |
| --- | --- | --- | --- | --- | --- |
| **Title and abstract** | | | | | |
|  | 1 | (a) Indicate the study’s design with a commonly used term in the title or the abstract (b) Provide in the abstract an informative and balanced summary of what was done and what was found | 1, 3 | RECORD 1.1: The type of data used should be specified in the title or abstract. When possible, the name of the databases used should be included.  RECORD 1.2: If applicable, the geographic region and timeframe within which the study took place should be reported in the title or abstract.  RECORD 1.3: If linkage between databases was conducted for the study, this should be clearly stated in the title or abstract. | 1,3 |
| **Introduction** | | | | | |
| Background rationale | 2 | Explain the scientific background and rationale for the investigation being reported | 4-5 | N/A |  |
| Objectives | 3 | State specific objectives, including any prespecified hypotheses | 4-5 | N/A |  |
| **Methods** | | | | | |
| Study Design | 4 | Present key elements of study design early in the paper | 5 | N/A |  |
| Setting | 5 | Describe the setting, locations, and relevant dates, including periods of recruitment, exposure, follow-up, and data collection | 5 | N/A |  |
| Participants | 6 | (a) Cohort study - Give the eligibility criteria, and the sources and methods of selection of participants. Describe methods of follow-up  (b) Cohort study - For matched studies, give matching criteria and number of exposed and unexposed | 5 | RECORD 6.1: The methods of study population selection (such as codes or algorithms used to identify subjects) should be listed in detail. If this is not possible, an explanation should be provided.  RECORD 6.2: Any validation studies of the codes or algorithms used to select the population should be referenced. If validation was conducted for this study and not published elsewhere, detailed methods and results should be provided.  RECORD 6.3: If the study involved linkage of databases, consider use of a flow diagram or other graphical display to demonstrate the data linkage process, including the number of individuals with linked data at each stage. | 5 |
| Variables | 7 | Clearly define all outcomes, exposures, predictors, potential confounders, and effect modifiers. Give diagnostic criteria, if applicable. | 5-6 | RECORD 7.1: A complete list of codes and algorithms used to classify exposures, outcomes, confounders, and effect modifiers should be provided. If these cannot be reported, an explanation should be provided. | 5-6 |
| Data sources/ measurement | 8 | For each variable of interest, give sources of data and details of methods of assessment (measurement).  Describe comparability of assessment methods if there is more than one group | 5-6 | N/A |  |
| Bias | 9 | Describe any efforts to address potential sources of bias | 6 | N/A |  |
| Study size | 10 | Explain how the study size was arrived at | 5 | N/A |  |
| Quantitative variables | 11 | Explain how quantitative variables were handled in the analyses. If applicable, describe which groupings were chosen, and why | 6 | N/A |  |
| Statistical methods | 12 | (a) Describe all statistical methods, including those used to control for confounding  (b) Describe any methods used to examine subgroups and interactions  (c) Explain how missing data were addressed  (d) Cohort study - If applicable, explain how loss to follow-up was addressed  (e) Describe any sensitivity analyses | 6 | N/A |  |
| Data access and cleaning methods |  | N/A |  | RECORD 12.1: Authors should describe the extent to which the investigators had access to the database population used to create the study population.  RECORD 12.2: Authors should provide information on the data cleaning methods used in the study. | 5-6; 14 |
| Linkage |  | N/A |  | RECORD 12.3: State whether the study included person-level, institutional-level, or other data linkage across two or more databases. The methods of linkage and methods of linkage quality evaluation should be provided. | 5 |
| **Results** | | | | | |
| Participants | 13 | (a) Report the numbers of individuals at each stage of the study (e.g., numbers potentially eligible, examined for eligibility, confirmed eligible, included in the study, completing follow-up, and analysed)  (b) Give reasons for non-participation at each stage.  (c) Consider use of a flow diagram | Supplement 2 | RECORD 13.1: Describe in detail the selection of the persons included in the study (i.e., study population selection) including filtering based on data quality, data availability and linkage. The selection of included persons can be described in the text and/or by means of the study flow diagram. | 5 |
| Descriptive data | 14 | (a) Give characteristics of study participants (e.g., demographic, clinical, social) and information on exposures and potential confounders  (b) Indicate the number of participants with missing data for each variable of interest  (c) Cohort study - summarise follow-up time (e.g., average and total amount) | 7 | N/A |  |
| Outcome data | 15 | Cohort study - Report numbers of outcome events or summary measures over time | 7 | N/A |  |
| Main results | 16 | (a) Give unadjusted estimates and, if applicable, confounder-adjusted estimates and their precision (e.g., 95% confidence interval). Make clear which confounders were adjusted for and why they were included  (b) Report category boundaries when continuous variables were categorized  (c) If relevant, consider translating estimates of relative risk into absolute risk for a meaningful time period | 9-11 | N/A |  |
| Other analyses | 17 | Report other analyses done—e.g., analyses of subgroups and interactions, and sensitivity analyses | 7-11 | N/A |  |
| **Discussion** | | | | | |
| Key results | 18 | Summarise key results with reference to study objectives | 11 | N/A |  |
| Limitations | 19 | Discuss limitations of the study, taking into account sources of potential bias or imprecision. Discuss both direction and magnitude of any potential bias | 13 | RECORD 19.1: Discuss the implications of using data that were not created or collected to answer the specific research question(s). Include discussion of misclassification bias, unmeasured confounding, missing data, and changing eligibility over time, as they pertain to the study being reported. | 11-13 |
| Interpretation | 20 | Give a cautious overall interpretation of results considering objectives, limitations, multiplicity of analyses, results from similar studies, and other relevant evidence | 11-13 | N/A |  |
| Generalisability | 21 | Discuss the generalisability (external validity) of the study results | 13 | N/A |  |
| **Other Information** | | | | | |
| Funding | 22 | Give the source of funding and the role of the funders for the present study and, if applicable, for the original study on which the present article is based | 14-15 | N/A |  |
| Accessibility of protocol, raw data, and programming code |  | N/A |  | RECORD 22.1: Authors should provide information on how to access any supplemental information such as the study protocol, raw data, or programming code. | 14 |

*Reference: Benchimol, E. I., Smeeth, L., Guttmann, A., Harron, K., Moher, D., Petersen, I., ... & RECORD Working Committee. (2015). The REporting of studies Conducted using Observational Routinely-collected health Data (RECORD) statement. PLoS Med, 12(10), e1001885.

*Checklist is protected under Creative Commons Attribution ([CC BY](http://creativecommons.org/licenses/by/4.0/)) license.

**Supplement 2 – Study flow diagram.**

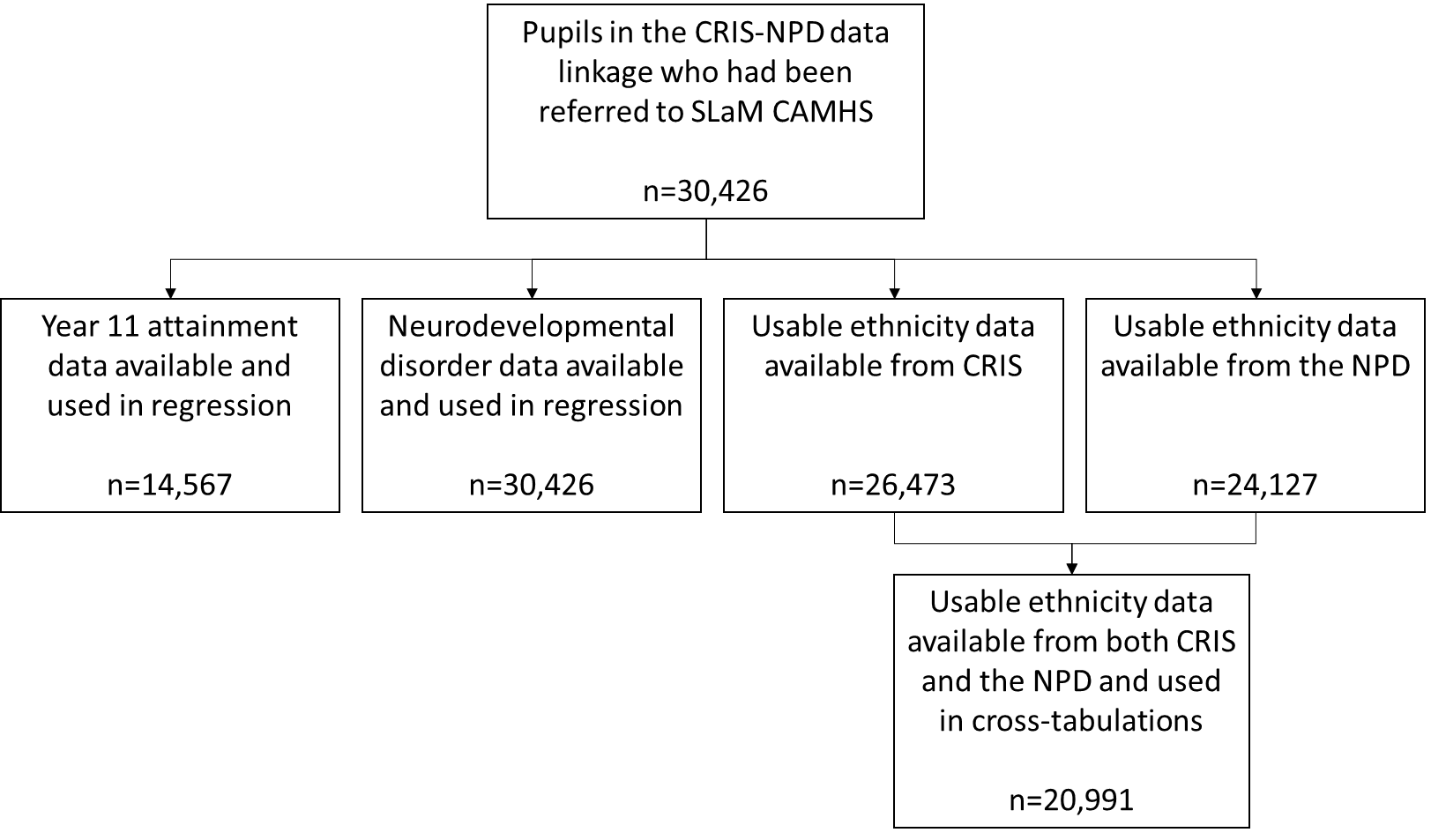

**Supplement 3 – Ethnic groups in the NPD.**

| **Approved extended categories** | **Minor ethnic group** | **Major ethnic group** |
| --- | --- | --- |
| White – British | White - British | White |
| White – Cornish ^a^ |  |  |
| White – English |  |  |
| White – Scottish |  |  |
| White – Welsh |  |  |
| Other White British |  |  |
| White – Irish | White |  |
| Traveller of Irish heritage |  |  |
| Any other White background | Any other White background |  |
| Albanian |  |  |
| Bosnian-Herzegovinian |  |  |
| Croatian |  |  |
| Greek/Greek Cypriot |  |  |
| Greek |  |  |
| Greek Cypriot |  |  |
| Italian ^a^ |  |  |
| Kosovan |  |  |
| Portuguese |  |  |
| Serbian |  |  |
| Turkish/Turkish Cypriot |  |  |
| Turkish |  |  |
| Turkish Cypriot |  |  |
| White European ^a^ |  |  |
| White Eastern European ^a^ |  |  |
| White Western European ^a^ |  |  |
| White other |  |  |
| Gypsy/Roma | Gypsy/Roma |  |
| Gypsy |  |  |
| Roma |  |  |
| Other Gypsy/Roma |  |  |
| White and Black Caribbean | White and Black Caribbean | Mixed/Dual background |
| White and Black African | White and Black African |  |
| White and Asian | White and Asian |  |
| White and Pakistani ^a^ |  |  |
| White and Indian ^a^ |  |  |
| White and any other Asian background |  |  |
| Any other Mixed background | Any other Mixed background |  |
| Asian and any other ethnic group ^a^ |  |  |
| Asian and Black |  |  |
| Asian and Chinese |  |  |
| Black and any other ethnic group ^a^ |  |  |
| Black and Chinese |  |  |
| Chinese and any other ethnic group ^a^ |  |  |
| White and any other ethnic group ^a^ |  |  |
| White and Chinese |  |  |
| Other Mixed background |  |  |
| Indian | Indian | Asian or Asian British |
| Pakistani | Pakistani |  |
| Mirpuri Pakistani ^a^ |  |  |
| Kashmiri Pakistani ^a^ |  |  |
| Other Pakistani |  |  |
| Bangladeshi | Bangladeshi |  |
| Any other Asian background | Any other Asian background |  |
| African Asian ^a^ |  |  |
| Kashmiri other ^a^ |  |  |
| Nepali ^a^ |  |  |
| Sri Lankan Sinhalese |  |  |
| Sri Lankan Tamil |  |  |
| Sri Lankan other |  |  |
| Other Asian |  |  |
| Black Caribbean | Black Caribbean | Black or Black British |
| Black - African | Black - African |  |
| Black - Angolan |  |  |
| Black - Congolese ^a^ |  |  |
| Black – Ghanaian |  |  |
| Black – Nigerian |  |  |
| Black - Sierra Leonean ^a^ |  |  |
| Black – Somali |  |  |
| Black – Sudanese |  |  |
| Other Black African |  |  |
| Any other Black background | Any other Black background |  |
| Black European ^a^ |  |  |
| Black North American ^a^ |  |  |
| Other Black |  |  |
| Chinese | Chinese | Chinese |
| Hong Kong Chinese ^a^ |  |  |
| Malaysian Chinese |  |  |
| Singaporean Chinese ^a^ |  |  |
| Taiwanese ^a^ |  |  |
| Other Chinese |  |  |
| Any other ethnic group | Any other ethnic group | Any other ethnic group |
| Afghan ^a^ |  |  |
| Arab other |  |  |
| Egyptian ^a^ |  |  |
| Filipino |  |  |
| Iranian |  |  |
| Iraqi |  |  |
| Japanese |  |  |
| Korean ^a^ |  |  |
| Kurdish |  |  |
| Latin/South/Central American ^a^ |  |  |
| Lebanese ^a^ |  |  |
| Libyan ^a^ |  |  |
| Malay |  |  |
| Moroccan ^a^ |  |  |
| Polynesian ^a^ |  |  |
| Thai ^a^ |  |  |
| Vietnamese |  |  |
| Yemeni |  |  |
| Other ethnic group |  |  |
| Refused | Refused | Refused |
| Information not yet obtained | Information not yet obtained | Information not yet obtained |

a. Specific ethnicities not included in the SLaM electronic health record ethnic categories list (CRIS, Supplement 4).

Note: Contains public sector information licensed under the Open Government Licence v3.0.

**Supplement 4 – Ethnic groups in CRIS.**

| **SLaM electronic health record ethnic categories** | **16+1 ethnic categories supplied from CRIS** |
| --- | --- |
| British (A) | British (A) |
| English (CA) |  |
| Scottish (CB) |  |
| Welsh (CC) |  |
| Irish (B) | Irish (B) |
| Irish Traveller (CL) |  |
| Albanian (CS) | Any other White background € |
| All former USSR Republics (CQ) ^a^ |  |
| Bosnian (CT) |  |
| Croatian (CU) |  |
| Cypriot (part not stated) (CE) ^a^ |  |
| Greek (CF) |  |
| Greek Cypriot (CG) |  |
| Gypsy/Romany (CN) |  |
| Kosovan (CR) |  |
| Kurdish (C5) |  |
| Other former Yugoslavia (CW) ^a^ |  |
| Other White Unspecified (C3) |  |
| Other White/Mixed European (CY) ^a^ |  |
| Portuguese (C4) |  |
| Serbian (CV) |  |
| Traveller (CM) ^a^ |  |
| Turkish (CH) |  |
| Turkish Cypriot (CJ) |  |
| White and Black Caribbean (D) | White and Black Caribbean (D) |
| White and Black African € | White and Black African € |
| White and Asian (F) | White and Asian (F) |
| Asian and Chinese (GE) | Any other Mixed background (G) |
| Black and Asian (GA) |  |
| Black and Chinese (GB) |  |
| Black and White (GC) ^a^ |  |
| Chinese and White (GD) |  |
| Indian/British Indian (H) | Indian (H) |
| Pakistani/British Pakistani (J) | Pakistani (J) |
| Bangladeshi/British Bangladeshi (K) | Bangladeshi (K) |
| British Asian (LH) ^a^ | Any other Asian background (L) |
| Caribbean Asian (LJ) ^a^ |  |
| East African Asian (LD) ^a^ |  |
| Mixed Asian (LA) ^a^ |  |
| Other Asian Unspecified (LK) |  |
| Sinhalese (LG) |  |
| Sri Lankan (LE) |  |
| Tamil (LF) |  |
| Caribbean (M) | Caribbean (M) |
| Algerian (PP) ^a^ | African (N) |
| Angolan (PJ) |  |
| Eritrean (PK) ^a^ |  |
| Ethiopian (PL) ^a^ |  |
| Ghanaian (PM) |  |
| Nigerian (PC) |  |
| Somali (PA) |  |
| Sudanese (PH) |  |
| Ugandan (PQ) ^a^ |  |
| Other African (N) |  |
| Mixed Black (PB) ^a^ | Any other Black background (P) |
| Black British (PD) ^a^ |  |
| Other Black Unspecified (PE) |  |
| Chinese (R) | Chinese (R) |
| Any Other Group (SE) | Any other ethnic group (S) |
| Arab (SG) |  |
| Columbian (SK) ^a^ |  |
| Ecuadorian (SL) ^a^ |  |
| Filipino (SC) |  |
| Iranian (SH) |  |
| Iraqi (SJ) |  |
| Japanese (SB) |  |
| Malaysian (SD) |  |
| Middle Eastern (SF) ^a^ |  |
| Other Latin American (SM) |  |
| Vietnamese (SA) |  |
| Not Stated (Z) | Not Stated (Z) |

a. Specific ethnicities not included in the ‘approved extended ethnic categories’ list (NPD, Supplement 3).

**Supplement 5 – Aggregating CRIS ethnicity to assist comparison with the NPD.**

| **16+1 ethnic categories supplied from CRIS** | **Aggregate category** |
| --- | --- |
| British (A) | White |
| Irish (B) |  |
| Any other White background (C) |  |
| White and Black Caribbean (D) | Mixed |
| White and Black African (E) |  |
| White and Asian (F) |  |
| Any other Mixed background (G) |  |
| Indian (H) | Asian |
| Pakistani (J) |  |
| Bangladeshi (K) |  |
| Any other Asian background (L) |  |
| Caribbean (M) | Black |
| African (N) |  |
| Any other Black background (P) |  |
| Chinese (R) | Chinese |
| Any other ethnic group (S) | Other |
| Not Stated (Z) | Unknown |

**Supplement 6 – Proportion of disaggregated CRIS ethnicities categorised as consistent major ethnic groups in the NPD, among individuals where ethnicity was available from both sources (n=20,991).**

|  |  | **NPD** | | | | | |
| --- | --- | --- | --- | --- | --- | --- | --- |
|  |  | **White**  **n=11,507** | **Black**  **n=5,532** | **Asian**  **n=632** | **Chinese**  **n=68** | **Mixed**  **n=2,810** | **Other**  **n=442** |
| **CRIS** | **British**  **n=10,634** | 10,056  (94.6%) |  |  |  |  |  |
|  | **Irish**  **n=129** | 121  (93.8%) |  |  |  |  |  |
|  | **Any other White background**  **n=1,090** | 877  (80.5%) |  |  |  |  |  |
|  | **African**  **n=1,216** |  | 1,114  (91.6%) |  |  |  |  |
|  | **Caribbean**  **n=1,207** |  | 1,089  (90.2%) |  |  |  |  |
|  | **Any other Black background**  **n=3,320** |  | 2,800  (84.3%) |  |  |  |  |
|  | **Indian**  **n=170** |  |  | 142  (83.5%) |  |  |  |
|  | **Pakistani**  **n=129** |  |  | 116  (89.9%) |  |  |  |
|  | **Bangladeshi**  **n=111** |  |  | 105  (94.6%) |  |  |  |
|  | **Any other Asian background**  **n=268** |  |  | 147  (54.9%) |  |  |  |
|  | **Chinese**  **n=58** |  |  |  | 51  (87.9%) |  |  |
|  | **White and Asian**  **n=134** |  |  |  |  | 104  (77.6%) |  |
|  | **White and Black African**  **n=238** |  |  |  |  | 179  (75.2%) |  |
|  | **White and Black Caribbean**  **n=1,102** |  |  |  |  | 873  (79.2%) |  |
|  | **Any other Mixed background**  **n=372** |  |  |  |  | 272  (73.1%) |  |
|  | **Any other ethnic group**  **n=813** |  |  |  |  |  | 216  (26.6%) |

Note: Row percentages provided. Off-diagonal proportions have been suppressed to avoid disclosive cell counts.
